# Procedural Volume and Outcomes with Atrial Fibrillation Ablation: A Report from the NCDR AFib Ablation Registry

**DOI:** 10.1101/2024.01.18.24301509

**Authors:** Sharma Kattel, Zhen Tan, Zhenqiu Lin, Reed Mszar, Prashanthan Sanders, Emily P. Zeitler, Paul C. Zei, T Jared Bunch, Moussa Mansour, Joseph Akar, Jeptha P. Curtis, Daniel J. Friedman, James V Freeman

## Abstract

**Background:** Hospital and physician procedure volumes have been associated with outcomes for several cardiac procedures; however, this has not been well defined for atrial fibrillation (AF) ablation in the contemporary practice.

**Objective:** To determine the association between hospital and physician procedural volume and success for AF ablation and the risk of major adverse events (MAE).

**Methods:** Procedures reported to the NCDR AFib Ablation Registry between July 2019 and June 2022 were included. Physician and hospital procedural volumes were annualized and stratified into quartiles (Q) to compare success and MAE. Three level hierarchical (patient, hospital and physician) generalized linear models were used to assess the adjusted relationship between procedural volume and the likelihood of procedural outcomes.

**Results:** A total of 70,296 first time AF ablation at 186 hospitals across the United States were included. The mean age was 66.1 ± 10.3 years, 36% were women, the mean CHA_2_DS_2_-VASc score was 2.7±1.6, and 57.4% of patients had paroxysmal AF. The median annual procedural volume for hospitals and physicians were 230 (IQR:144.7-307.2) and 66 (IQR: 44.7-97.5), respectively. Overall, acute procedural success was 98.5 % and the MAE rate was 1.0%. With Q4 (highest) hospital volume as a reference, the adjusted likelihood of procedural success was significantly lower for Q1 (OR: 0.44, CI: 0.29-0.68), Q2 (OR: 0.50, CI: 0.33-0.75) and Q3 (OR: 0.60, CI: 0.40-0.89). Similarly, for physician, procedural success was less among Q1 (OR: 0.38, CI: 0.28-0.51), Q2 (OR: 0.51, CI: 0.38-0.69) and Q3 (OR: 0.55, CI: 0.42-0.72). With MAE, compared with Q4, there was an inverse relationship between procedural volume for hospitals in Q1 (OR: 1.78, CI: 1.26-2.51) but not Q2 (OR: 1.06, CI: 0.77-1.46) or Q3 (OR: 1.19, CI: 0.89-1.58) and for physicians in Q1 (OR: 1.93, CI: 1.44-2.58) and Q2 (OR: 1.49, CI: 1.13-1.97) but not in Q3 (OR: 1.22, CI: 0.94-1.58). An adjusted MAE ≤ 1% was predicted by an annual volume of approximately 190 for hospitals and 60 for physicians.

**Conclusion:** In this national cohort, hospital and physician AF ablation procedural volumes were directly related to acute procedural success and inversely related to rates of MAE.

**Condensed Abstract:** The relationship between atrial fibrillation (AF) ablation procedure volume and outcomes is not well defined in contemporary practice. In this national cohort of 70,296 patients undergoing first time AF ablation, acute procedural success was relatively high (98.5%) and risk of major adverse event remained low (1.0%). Greater hospital and physician procedure volume was associated with higher procedural success. There was an inverse relationship between hospital and physician procedural volume and the risk of major adverse events.

## Introduction

Atrial fibrillation (AF) is one of the most common supraventricular arrhythmias worldwide^1,2^. Catheter ablation for AF has been associated with decreased arrhythmia burden, fewer symptoms, improved quality of life, and improved mortality in patients with reduced left ventricular function compared with medical therapy^3^. As a result, catheter ablation for AF has become one of the most commonly performed cardiovascular procedures ^3,4^.

For many cardiac surgical and percutaneous procedures including coronary artery bypass grafting^5^, mitral valve surgery ^6^, percutaneous coronary artery intervention ^7,8^, implantable cardioverter defibrillator implantation ^9^, and transcatheter aortic valve replacement ^10^, higher procedural volumes have been associated with improved outcomes. Despite these known associations between procedural volume and outcomes in other cardiac procedures, the relationship between physician and hospital procedure volumes and outcomes for AF catheter ablation in contemporary practice is not well studied. Prior studies evaluating this relationship had substantial limitations ^11,12^. These studies utilized data from the National Inpatient Sample (NIS), an administrative claims dataset of inpatient admissions in the US, however, the majority of AF ablations in contemporary practice are performed as outpatient procedures. These studies are therefore not generalizable, and are likely to overestimate procedural complications. In addition, these studies used older data from almost a decade ago, prior to significant increase in AF ablation volume and advances in AF ablation technologies that have substantially improved the safety profile of AF ablation. A recent meta-analysis of randomized controlled trials published between 2013-2022 showed a decreasing rate of complications with AF ablation but contemporary real-world data are lacking ^13^. Thus, it is not clear whether procedural volume significantly impacts the procedural safety and effectiveness of AF ablation in the current era. To address these knowledge gaps, we used data from the National Cardiovascular Data Registry (NCDR) AFib Ablation Registry to evaluate contemporary rates of procedural success and adverse events (AE) and the association between procedure volume and outcomes.

## Methods

This study was approved by the NCDR EP Suite Research and Publications Committee for data abstraction and analysis. Study analyses were performed by the Yale Center for Outcomes Research and Evaluation (CORE) Data Analytic Center and the study was approved by the Yale University Institutional Review Board.

### Data Source

The data for this study were derived from the NCDR AFib Ablation Registry, the largest prospective AF ablation registry in the world. The registry has been previously described ^14^ and the data collection methodology and quality of collected data have been described elsewhere ^15^. In brief, the registry is a large, nationwide multicenter registry initiated in 2016 to assess the demographics, management, and outcomes in patients undergoing percutaneous catheter ablation for AF. Participation in the registry is voluntary. Participating hospitals in the Registry are required to submit patient and procedure-related data on a quarterly basis to the American College of Cardiology Foundation’s NCDR database. A link to the full data collection forms for the index hospitalization is publicly available (https://cvquality.acc.org/NCDR-Home/data-collection)^16^. The NCDR Data Quality Reporting (DQR) process is designed to ensure that submissions are complete, valid, and accurate; it involves an annual audit of approximately 5% of the participating sites that are randomly selected for submitted data comparison to source documentation and billing data ^15^.

### Study Population and Annualized Procedural Volume

We included patients ≥18 years of age enrolled in the NCDR AFib Ablation Registry undergoing AF ablation between July 1, 2019 to June 30, 2022. We limited the study to this time period to assess the recent trends of procedural success and AE in AF ablation as there has been substantial improvement in ablation catheter designs, electro-anatomical mapping techniques and changes in practice including regular use of intracardiac echocardiogram. We excluded patients with prior surgical or catheter ablation for AF and those with missing data regarding pulmonary vein isolation. The cohort was stratified into quartiles of hospital and physician volume for all subsequent analyses.

We annualized hospital and physician procedure volumes for the hospitals and physicians. Annualized hospital and physician procedure volumes were calculated by tabulating the total number of procedures performed at a hospital or by a physician in the last 3 years, divided by the number of months during the study period and multiplying the number by 12. Hospitals and physicians were then separately stratified into quartiles of annualized procedure volume.

### Study Endpoints

The main outcomes were assessed as safety and effectiveness endpoints. The primary safety outcome was any MAE and the secondary safety outcome was any AE. The primary effectiveness endpoint was acute procedural success, defined as complete isolation of all pulmonary veins at the time of ablation. MAE was defined as a composite of death, stroke, TIA, cardiac arrest, acute myocardial infraction, pericardial effusion requiring intervention, major bleeding, and vascular injury requiring intervention. Any AE included MAE and any other in-hospital AE documented in the registry data collection forms^16^.

### Statistical Analysis

The baseline clinical, procedural, hospital characteristics, safety and effectiveness endpoints were compared among quartiles of annualized hospital and physician procedure volumes separately using the Wilcoxon rank sum test for continuous variables and the χ^2^ test for categorical variables.

Logistic regression was used to compare the relationship between volume quartiles and the likelihood of procedural outcomes compared with the highest volume quartile (Q4). Three level hierarchical (patient, hospital, and physician) generalized linear models were used to assess the adjusted relationship between procedural volume and the likelihood of procedural outcomes. To facilitate the model convergence, patient level was summarized to a logit score derived from the patients’ clinical characteristics associated with the likelihood of an outcome and clinical relevance. Variables used for generation of the logit score were those known to be associated with the study outcomes and included: age, sex, BMI, GFR, left ventricular systolic dysfunction, CHF, cardiomyopathy, hypertension, diabetes, prior stroke or TIA, prior thromboembolic event, vascular disease, coronary artery disease, chronic lung disease, sleep apnea, valvular heart disease, and prior attempts to restore sinus rhythm.

The relationship between procedural volume and outcomes was assessed using generalized linear mixed models. Adjusted and unadjusted models were created. Quadratic regression was used to examine the potentially nonlinear relationship between hospital or physician volume and outcome.

A sensitivity analysis was performed involving all AF ablation cases (de-novo and re-do ablation) to assess if there was any change in procedure volume outcome when re-do AF ablations were included. A secondary adjusted analysis was performed that included the total procedural volume performed by a hospital or a physician during the study period to assess if cumulative exposure impacted the annualized volume outcome relationship.

A p-value of <0.05 was considered statistically significant. Analyses were performed using SAS (version 9.4, SAS institute, Cary, North Carolina).

Mr. Tan and Drs. Kattel and Freeman had full access to all data in the study and take responsibility for the integrity of the data and the accuracy of the data analysis.

## Results

A total of 70,296 patients who underwent first time AF ablation in 186 hospitals across the US, involving 753 individual physicians were included (**Figure 1**).

**Figure 1:**
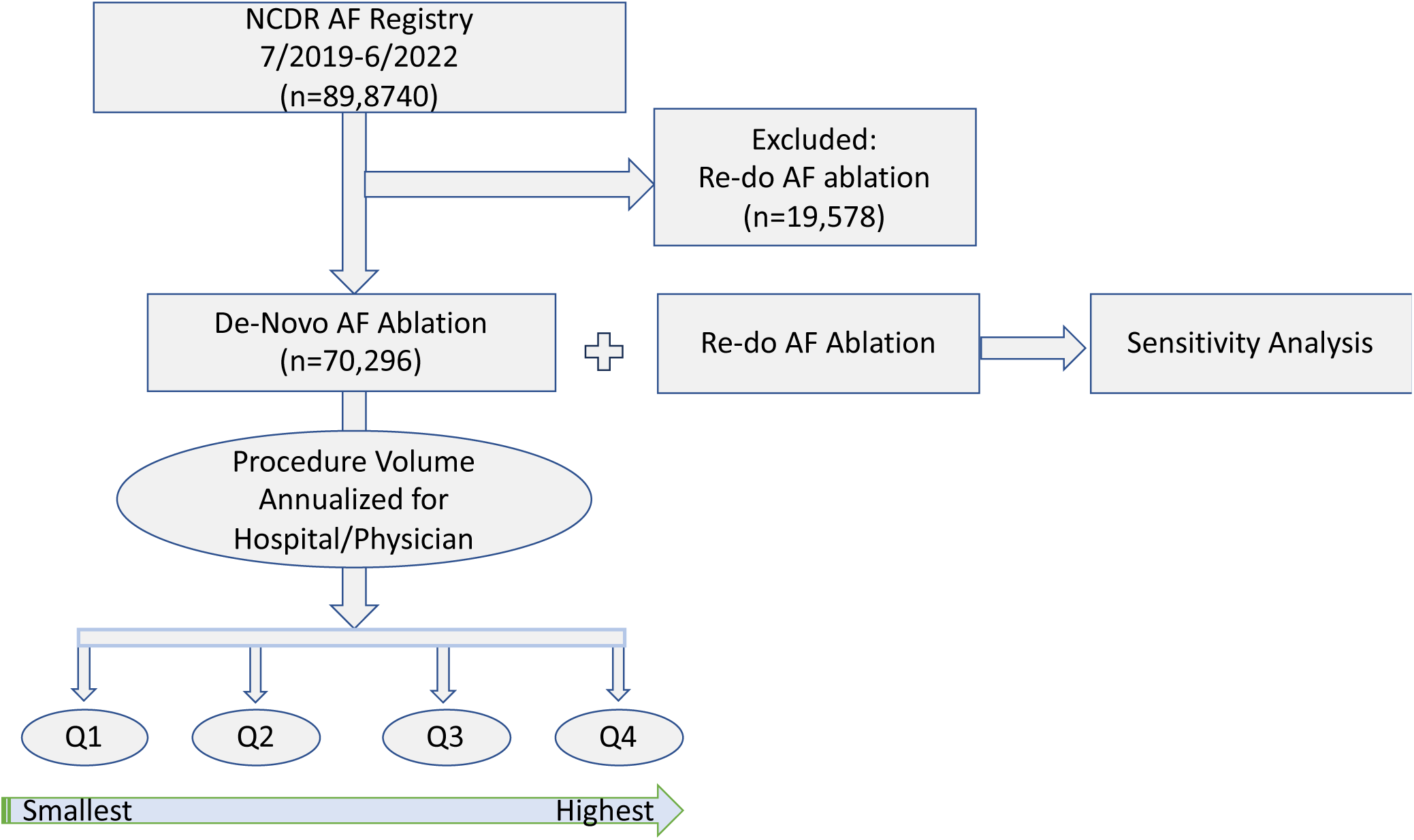
**Study Overview**. NCDR, National Cardiovascular Data Registry; AF, Atrial Fibrillation; Q, Procedure Volume Quartiles

### Baseline Patient and Hospital Characteristics

The baseline demographics, clinical characteristics, and procedural outcomes are depicted in **Tables 1, 2, and 3**. In the overall cohort, the mean age was 66.1 ± 10.3 years, most were white (92.1%), 36.0% were female, and mean CHA_2_DS_2_-VASc score was 2.7 ± 1.6. The 57.4% were paroxysmal AF and 37.2% were prescribed an antiarrhythmic drug at the time of ablation. Most procedures were performed in urban (65.1%) or suburban (29.4%) hospitals.

**Table 1.**
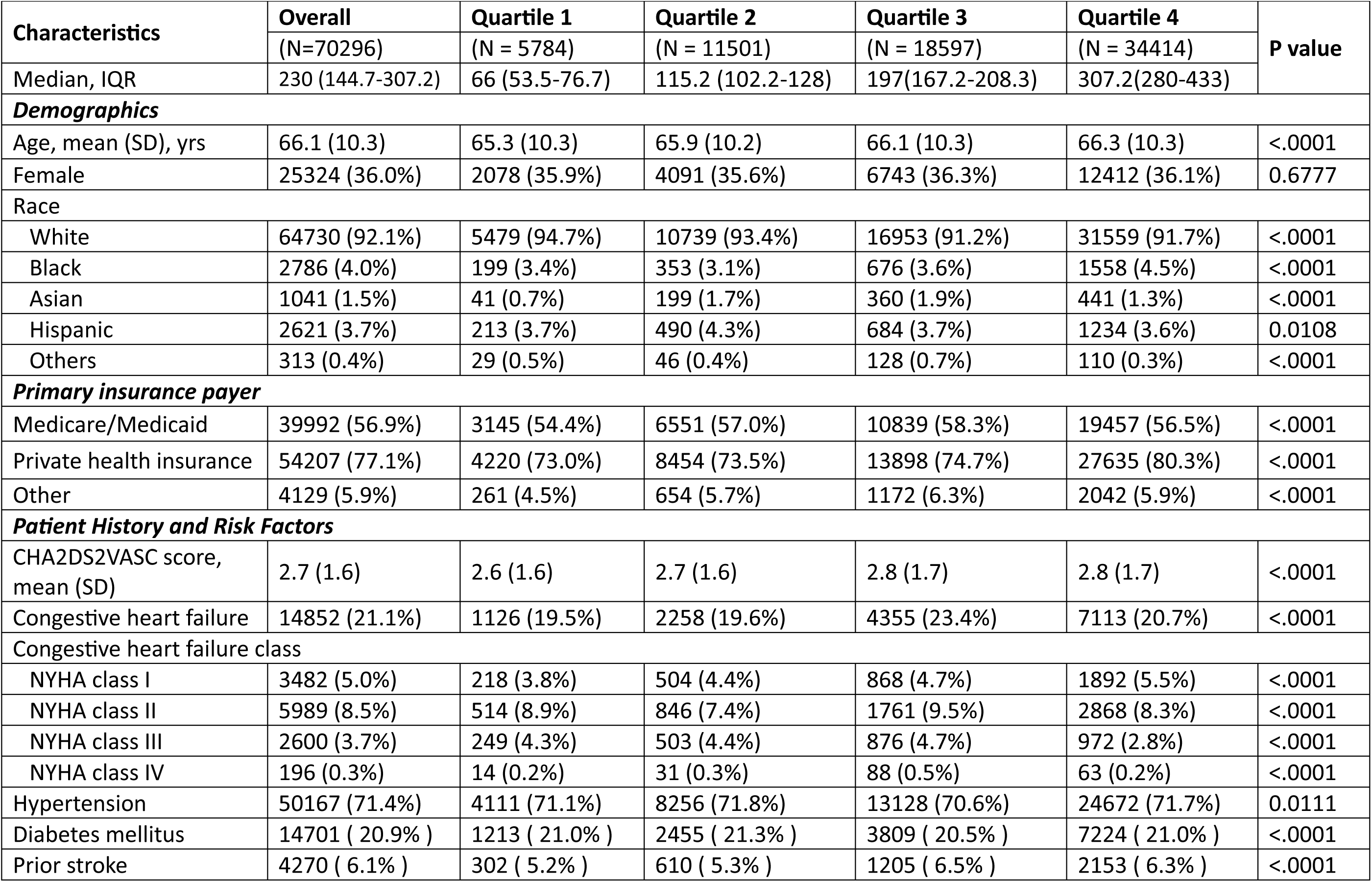
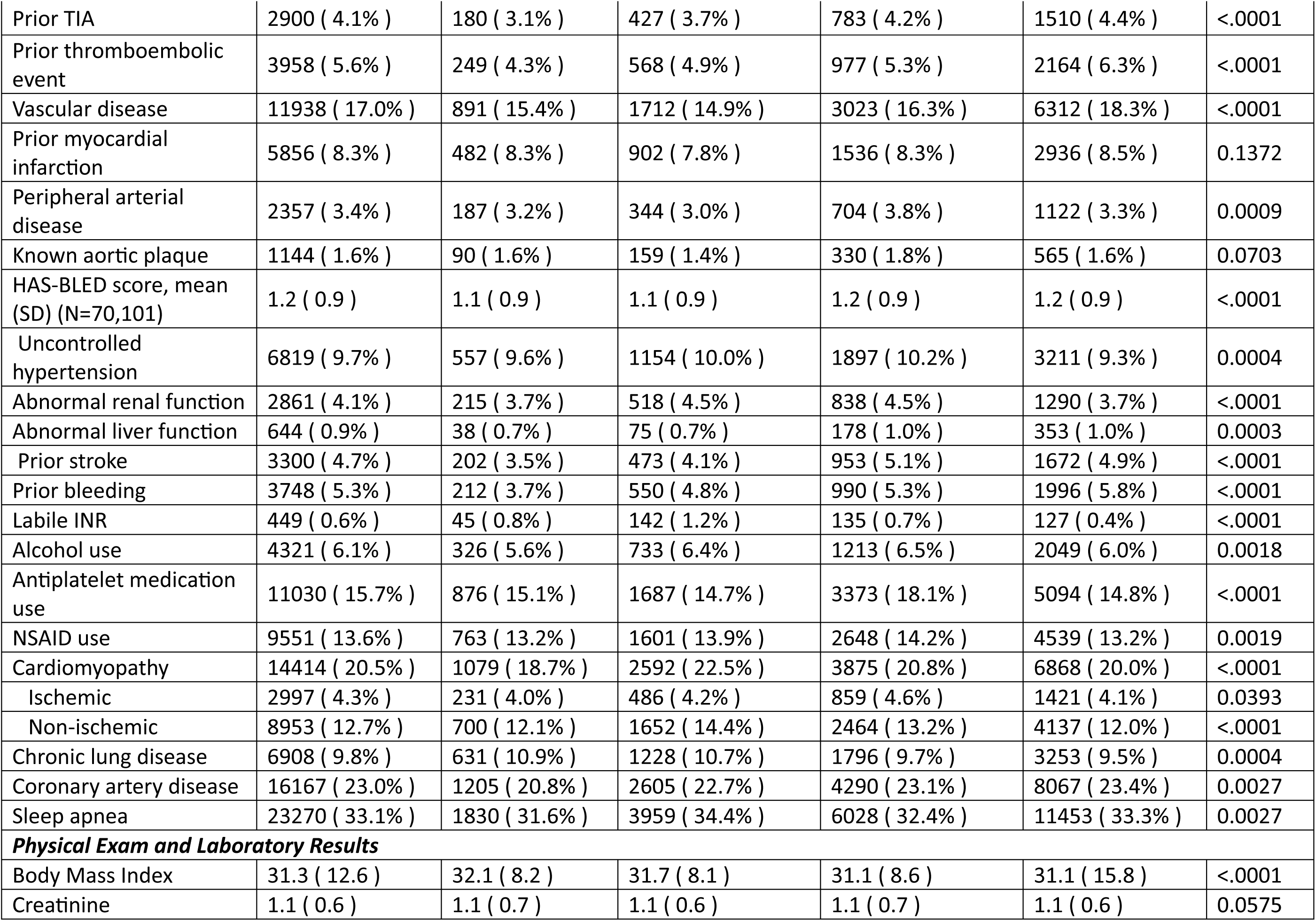
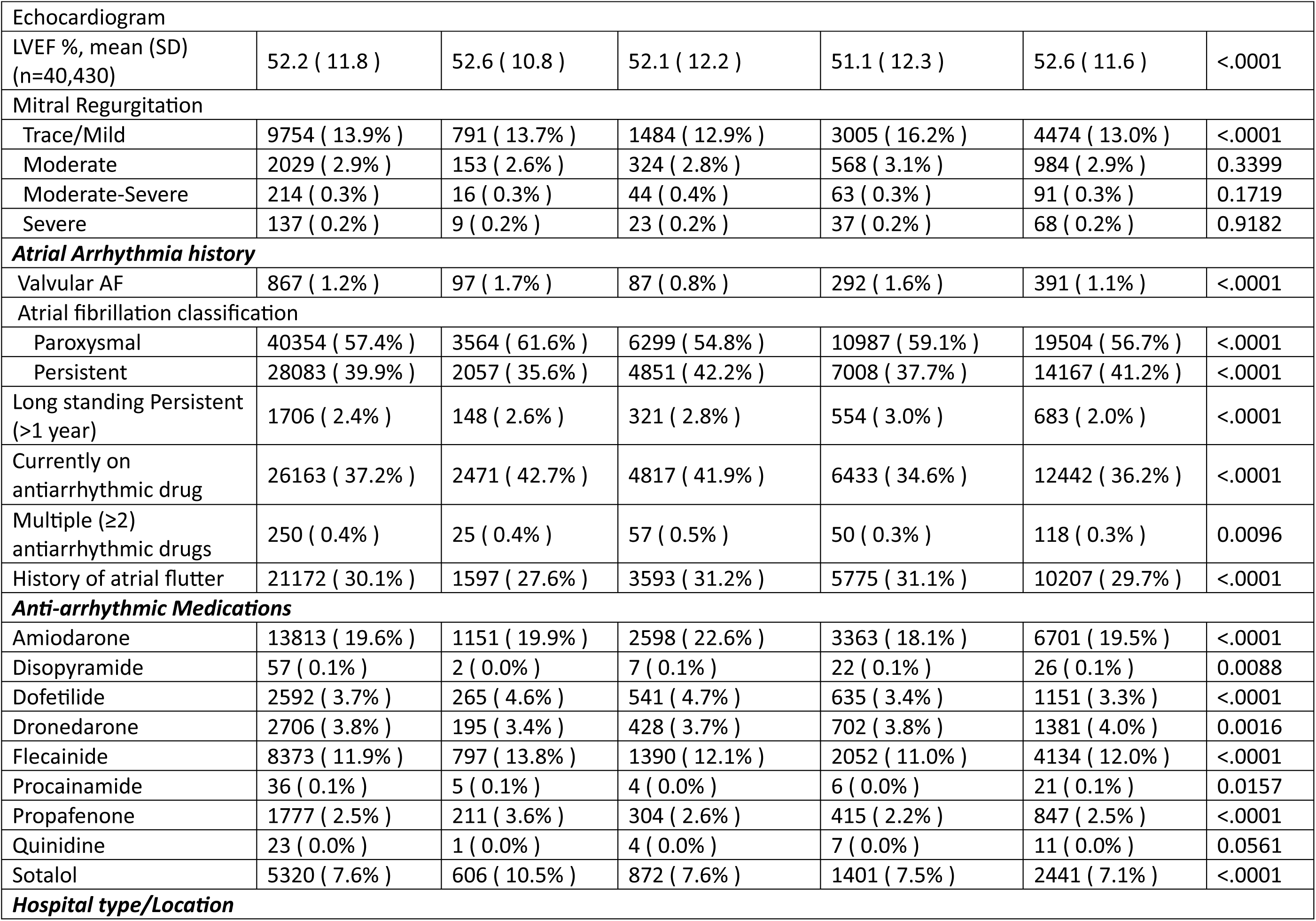
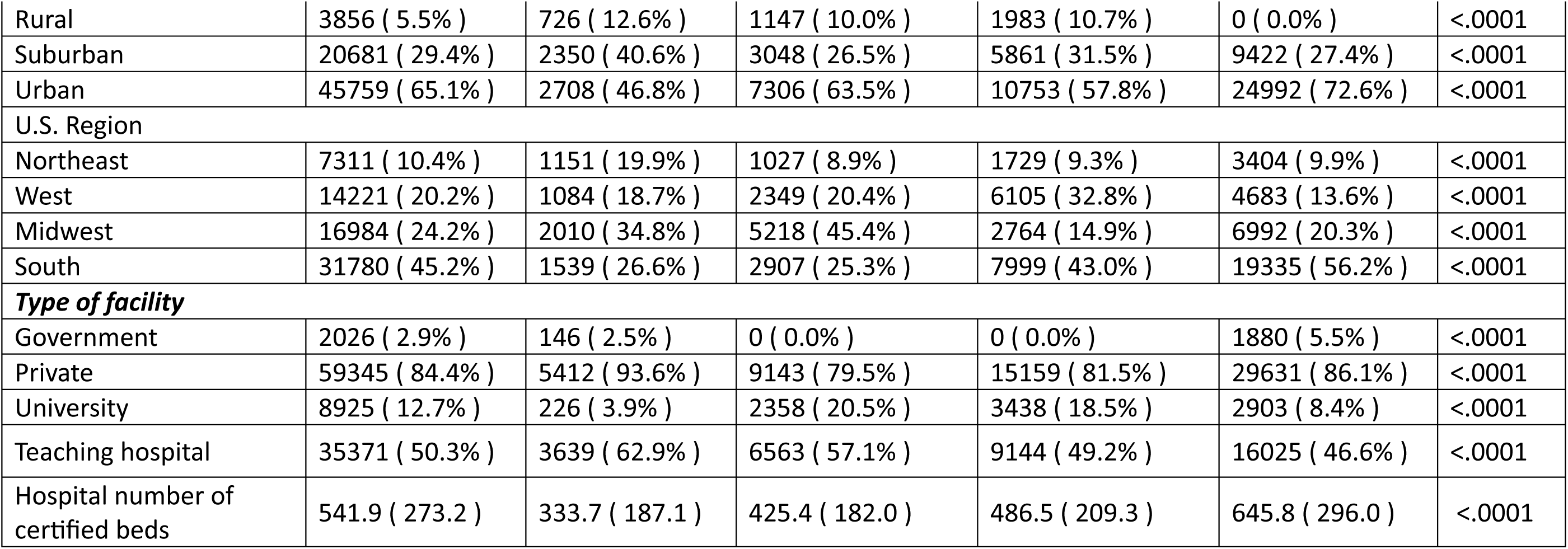
Patient Characteristics Stratified by Quartile of Annualized Hospital Volume in Atrial Fibrillation Ablation.

|  |  |  |  |  |  |  |
| --- | --- | --- | --- | --- | --- | --- |
| **Characteristics** | **Overall** | **Quartile 1** | **Quartile 2** | **Quartile 3** | **Quartile 4** | **P value** |
|  | (N=70296) | (N = 5784) | (N = 11501) | (N = 18597) | (N = 34414) |  |
| Median, IQR | 230 (144.7-307.2) | 66 (53.5-76.7) | 115.2 (102.2-128) | 197(167.2-208.3) | 307.2(280-433) |  |
| ***Demographics*** |  |  |  |  |  |  |
| Age, mean (SD), yrs | 66.1 (10.3) | 65.3 (10.3) | 65.9 (10.2) | 66.1 (10.3) | 66.3 (10.3) | <.0001 |
| Female | 25324 (36.0%) | 2078 (35.9%) | 4091 (35.6%) | 6743 (36.3%) | 12412 (36.1%) | 0.6777 |
| **Race** |  |  |  |  |  |  |
| White | 64730 (92.1%) | 5479 (94.7%) | 10739 (93.4%) | 16953 (91.2%) | 31559 (91.7%) | <.0001 |
| Black | 2786 (4.0%) | 199 (3.4%) | 353 (3.1%) | 676 (3.6%) | 1558 (4.5%) | <.0001 |
| Asian | 1041 (1.5%) | 41 (0.7%) | 199 (1.7%) | 360 (1.9%) | 441 (1.3%) | <.0001 |
| Hispanic | 2621 (3.7%) | 213 (3.7%) | 490 (4.3%) | 684 (3.7%) | 1234 (3.6%) | 0.0108 |
| Others | 313 (0.4%) | 29 (0.5%) | 46 (0.4%) | 128 (0.7%) | 110 (0.3%) | <.0001 |
| ***Primary insurance payer*** |  |  |  |  |  |  |
| Medicare/Medicaid | 39992 (56.9%) | 3145 (54.4%) | 6551 (57.0%) | 10839 (58.3%) | 19457 (56.5%) | <.0001 |
| Private health insurance | 54207 (77.1%) | 4220 (73.0%) | 8454 (73.5%) | 13898 (74.7%) | 27635 (80.3%) | <.0001 |
| Other | 4129 (5.9%) | 261 (4.5%) | 654 (5.7%) | 1172 (6.3%) | 2042 (5.9%) | <.0001 |
| ***Patient History and Risk Factors*** |  |  |  |  |  |  |
| CHA2DS2VASC score, mean (SD) | 2.7 (1.6) | 2.6 (1.6) | 2.7 (1.6) | 2.8 (1.7) | 2.8 (1.7) | <.0001 |
| Congestive heart failure | 14852 (21.1%) | 1126 (19.5%) | 2258 (19.6%) | 4355 (23.4%) | 7113 (20.7%) | <.0001 |
| **Congestive heart failure class** |  |  |  |  |  |  |
| NYHA class I | 3482 (5.0%) | 218 (3.8%) | 504 (4.4%) | 868 (4.7%) | 1892 (5.5%) | <.0001 |
| NYHA class II | 5989 (8.5%) | 514 (8.9%) | 846 (7.4%) | 1761 (9.5%) | 2868 (8.3%) | <.0001 |
| NYHA class III | 2600 (3.7%) | 249 (4.3%) | 503 (4.4%) | 876 (4.7%) | 972 (2.8%) | <.0001 |
| NYHA class IV | 196 (0.3%) | 14 (0.2%) | 31 (0.3%) | 88 (0.5%) | 63 (0.2%) | <.0001 |
| Hypertension | 50167 (71.4%) | 4111 (71.1%) | 8256 (71.8%) | 13128 (70.6%) | 24672 (71.7%) | 0.0111 |
| Diabetes mellitus | 14701 ( 20.9% ) | 1213 ( 21.0% ) | 2455 ( 21.3% ) | 3809 ( 20.5% ) | 7224 ( 21.0% ) | <.0001 |
| Prior stroke | 4270 ( 6.1% ) | 302 ( 5.2% ) | 610 ( 5.3% ) | 1205 ( 6.5% ) | 2153 ( 6.3% ) | <.0001 |
| Prior TIA | 2900 ( 4.1% ) | 180 ( 3.1% ) | 427 ( 3.7% ) | 783 ( 4.2% ) | 1510 ( 4.4% ) | <.0001 |
| Prior thromboembolic event | 3958 ( 5.6% ) | 249 ( 4.3% ) | 568 ( 4.9% ) | 977 ( 5.3% ) | 2164 ( 6.3% ) | <.0001 |
| Vascular disease | 11938 ( 17.0% ) | 891 ( 15.4% ) | 1712 ( 14.9% ) | 3023 ( 16.3% ) | 6312 ( 18.3% ) | <.0001 |
| Prior myocardial infarction | 5856 ( 8.3% ) | 482 ( 8.3% ) | 902 ( 7.8% ) | 1536 ( 8.3% ) | 2936 ( 8.5% ) | 0.1372 |
| Peripheral arterial disease | 2357 ( 3.4% ) | 187 ( 3.2% ) | 344 ( 3.0% ) | 704 ( 3.8% ) | 1122 ( 3.3% ) | 0.0009 |
| Known aortic plaque | 1144 ( 1.6% ) | 90 ( 1.6% ) | 159 ( 1.4% ) | 330 ( 1.8% ) | 565 ( 1.6% ) | 0.0703 |
| HAS-BLED score, mean (SD) (N=70,101) | 1.2 ( 0.9 ) | 1.1 ( 0.9 ) | 1.1 ( 0.9 ) | 1.2 ( 0.9 ) | 1.2 ( 0.9 ) | <.0001 |
| Uncontrolled hypertension | 6819 ( 9.7% ) | 557 ( 9.6% ) | 1154 ( 10.0% ) | 1897 ( 10.2% ) | 3211 ( 9.3% ) | 0.0004 |
| Abnormal renal function | 2861 ( 4.1% ) | 215 ( 3.7% ) | 518 ( 4.5% ) | 838 ( 4.5% ) | 1290 ( 3.7% ) | <.0001 |
| Abnormal liver function | 644 ( 0.9% ) | 38 ( 0.7% ) | 75 ( 0.7% ) | 178 ( 1.0% ) | 353 ( 1.0% ) | 0.0003 |
| Prior stroke | 3300 ( 4.7% ) | 202 ( 3.5% ) | 473 ( 4.1% ) | 953 ( 5.1% ) | 1672 ( 4.9% ) | <.0001 |
| Prior bleeding | 3748 ( 5.3% ) | 212 ( 3.7% ) | 550 ( 4.8% ) | 990 ( 5.3% ) | 1996 ( 5.8% ) | <.0001 |
| Labile INR | 449 ( 0.6% ) | 45 ( 0.8% ) | 142 ( 1.2% ) | 135 ( 0.7% ) | 127 ( 0.4% ) | <.0001 |
| Alcohol use | 4321 ( 6.1% ) | 326 ( 5.6% ) | 733 ( 6.4% ) | 1213 ( 6.5% ) | 2049 ( 6.0% ) | 0.0018 |
| Antiplatelet medication use | 11030 ( 15.7% ) | 876 ( 15.1% ) | 1687 ( 14.7% ) | 3373 ( 18.1% ) | 5094 ( 14.8% ) | <.0001 |
| NSAID use | 9551 ( 13.6% ) | 763 ( 13.2% ) | 1601 ( 13.9% ) | 2648 ( 14.2% ) | 4539 ( 13.2% ) | 0.0019 |
| Cardiomyopathy | 14414 ( 20.5% ) | 1079 ( 18.7% ) | 2592 ( 22.5% ) | 3875 ( 20.8% ) | 6868 ( 20.0% ) | <.0001 |
| Ischemic | 2997 ( 4.3% ) | 231 ( 4.0% ) | 486 ( 4.2% ) | 859 ( 4.6% ) | 1421 ( 4.1% ) | 0.0393 |
| Non-ischemic | 8953 ( 12.7% ) | 700 ( 12.1% ) | 1652 ( 14.4% ) | 2464 ( 13.2% ) | 4137 ( 12.0% ) | <.0001 |
| Chronic lung disease | 6908 ( 9.8% ) | 631 ( 10.9% ) | 1228 ( 10.7% ) | 1796 ( 9.7% ) | 3253 ( 9.5% ) | 0.0004 |
| Coronary artery disease | 16167 ( 23.0% ) | 1205 ( 20.8% ) | 2605 ( 22.7% ) | 4290 ( 23.1% ) | 8067 ( 23.4% ) | 0.0027 |
| Sleep apnea | 23270 ( 33.1% ) | 1830 ( 31.6% ) | 3959 ( 34.4% ) | 6028 ( 32.4% ) | 11453 ( 33.3% ) | 0.0027 |
| **Physical Exam and Laboratory Results** |  |  |  |  |  |  |
| Body Mass Index | 31.3 ( 12.6 ) | 32.1 ( 8.2 ) | 31.7 ( 8.1 ) | 31.1 ( 8.6 ) | 31.1 ( 15.8 ) | <.0001 |
| Creatinine | 1.1 ( 0.6 ) | 1.1 ( 0.7 ) | 1.1 ( 0.6 ) | 1.1 ( 0.7 ) | 1.1 ( 0.6 ) | 0.0575 |
| **Echocardiogram** |  |  |  |  |  |  |
| LVEF %, mean (SD)<br>(n=40,430) | 52.2 ( 11.8 ) | 52.6 ( 10.8 ) | 52.1 ( 12.2 ) | 51.1 ( 12.3 ) | 52.6 ( 11.6 ) | <.0001 |
| **Mitral Regurgitation** |  |  |  |  |  |  |
| Trace/Mild | 9754 ( 13.9% ) | 791 ( 13.7% ) | 1484 ( 12.9% ) | 3005 ( 16.2% ) | 4474 ( 13.0% ) | <.0001 |
| Moderate | 2029 ( 2.9% ) | 153 ( 2.6% ) | 324 ( 2.8% ) | 568 ( 3.1% ) | 984 ( 2.9% ) | 0.3399 |
| Moderate-Severe | 214 ( 0.3% ) | 16 ( 0.3% ) | 44 ( 0.4% ) | 63 ( 0.3% ) | 91 ( 0.3% ) | 0.1719 |
| Severe | 137 ( 0.2% ) | 9 ( 0.2% ) | 23 ( 0.2% ) | 37 ( 0.2% ) | 68 ( 0.2% ) | 0.9182 |
| **Atrial Arrhythmia history** |  |  |  |  |  |  |
| Valvular AF | 867 ( 1.2% ) | 97 ( 1.7% ) | 87 ( 0.8% ) | 292 ( 1.6% ) | 391 ( 1.1% ) | <.0001 |
| **Atrial fibrillation classification** |  |  |  |  |  |  |
| Paroxysmal | 40354 ( 57.4% ) | 3564 ( 61.6% ) | 6299 ( 54.8% ) | 10987 ( 59.1% ) | 19504 ( 56.7% ) | <.0001 |
| Persistent | 28083 ( 39.9% ) | 2057 ( 35.6% ) | 4851 ( 42.2% ) | 7008 ( 37.7% ) | 14167 ( 41.2% ) | <.0001 |
| Long standing Persistent<br>(>1 year) | 1706 ( 2.4% ) | 148 ( 2.6% ) | 321 ( 2.8% ) | 554 ( 3.0% ) | 683 ( 2.0% ) | <.0001 |
| Currently on<br>antiarrhythmic drug | 26163 ( 37.2% ) | 2471 ( 42.7% ) | 4817 ( 41.9% ) | 6433 ( 34.6% ) | 12442 ( 36.2% ) | <.0001 |
| Multiple (≥2)<br>antiarrhythmic drugs | 250 ( 0.4% ) | 25 ( 0.4% ) | 57 ( 0.5% ) | 50 ( 0.3% ) | 118 ( 0.3% ) | 0.0096 |
| History of atrial flutter | 21172 ( 30.1% ) | 1597 ( 27.6% ) | 3593 ( 31.2% ) | 5775 ( 31.1% ) | 10207 ( 29.7% ) | <.0001 |
| **Anti-arrhythmic Medications** |  |  |  |  |  |  |
| Amiodarone | 13813 ( 19.6% ) | 1151 ( 19.9% ) | 2598 ( 22.6% ) | 3363 ( 18.1% ) | 6701 ( 19.5% ) | <.0001 |
| Disopyramide | 57 ( 0.1% ) | 2 ( 0.0% ) | 7 ( 0.1% ) | 22 ( 0.1% ) | 26 ( 0.1% ) | 0.0088 |
| Dofetilide | 2592 ( 3.7% ) | 265 ( 4.6% ) | 541 ( 4.7% ) | 635 ( 3.4% ) | 1151 ( 3.3% ) | <.0001 |
| Dronedarone | 2706 ( 3.8% ) | 195 ( 3.4% ) | 428 ( 3.7% ) | 702 ( 3.8% ) | 1381 ( 4.0% ) | 0.0016 |
| Flecainide | 8373 ( 11.9% ) | 797 ( 13.8% ) | 1390 ( 12.1% ) | 2052 ( 11.0% ) | 4134 ( 12.0% ) | <.0001 |
| Procainamide | 36 ( 0.1% ) | 5 ( 0.1% ) | 4 ( 0.0% ) | 6 ( 0.0% ) | 21 ( 0.1% ) | 0.0157 |
| Propafenone | 1777 ( 2.5% ) | 211 ( 3.6% ) | 304 ( 2.6% ) | 415 ( 2.2% ) | 847 ( 2.5% ) | <.0001 |
| Quinidine | 23 ( 0.0% ) | 1 ( 0.0% ) | 4 ( 0.0% ) | 7 ( 0.0% ) | 11 ( 0.0% ) | 0.0561 |
| Sotalol | 5320 ( 7.6% ) | 606 ( 10.5% ) | 872 ( 7.6% ) | 1401 ( 7.5% ) | 2441 ( 7.1% ) | <.0001 |
| **Hospital type/Location** |  |  |  |  |  |  |
| Rural | 3856 ( 5.5% ) | 726 ( 12.6% ) | 1147 ( 10.0% ) | 1983 ( 10.7% ) | 0 ( 0.0% ) | <.0001 |
| Suburban | 20681 ( 29.4% ) | 2350 ( 40.6% ) | 3048 ( 26.5% ) | 5861 ( 31.5% ) | 9422 ( 27.4% ) | <.0001 |
| Urban | 45759 ( 65.1% ) | 2708 ( 46.8% ) | 7306 ( 63.5% ) | 10753 ( 57.8% ) | 24992 ( 72.6% ) | <.0001 |
| U.S. Region |  |  |  |  |  |  |
| Northeast | 7311 ( 10.4% ) | 1151 ( 19.9% ) | 1027 ( 8.9% ) | 1729 ( 9.3% ) | 3404 ( 9.9% ) | <.0001 |
| West | 14221 ( 20.2% ) | 1084 ( 18.7% ) | 2349 ( 20.4% ) | 6105 ( 32.8% ) | 4683 ( 13.6% ) | <.0001 |
| Midwest | 16984 ( 24.2% ) | 2010 ( 34.8% ) | 5218 ( 45.4% ) | 2764 ( 14.9% ) | 6992 ( 20.3% ) | <.0001 |
| South | 31780 ( 45.2% ) | 1539 ( 26.6% ) | 2907 ( 25.3% ) | 7999 ( 43.0% ) | 19335 ( 56.2% ) | <.0001 |
| **Type of facility** |  |  |  |  |  |  |
| Government | 2026 ( 2.9% ) | 146 ( 2.5% ) | 0 ( 0.0% ) | 0 ( 0.0% ) | 1880 ( 5.5% ) | <.0001 |
| Private | 59345 ( 84.4% ) | 5412 ( 93.6% ) | 9143 ( 79.5% ) | 15159 ( 81.5% ) | 29631 ( 86.1% ) | <.0001 |
| University | 8925 ( 12.7% ) | 226 ( 3.9% ) | 2358 ( 20.5% ) | 3438 ( 18.5% ) | 2903 ( 8.4% ) | <.0001 |
| Teaching hospital | 35371 ( 50.3% ) | 3639 ( 62.9% ) | 6563 ( 57.1% ) | 9144 ( 49.2% ) | 16025 ( 46.6% ) | <.0001 |
| Hospital number of certified beds | 541.9 ( 273.2 ) | 333.7 ( 187.1 ) | 425.4 ( 182.0 ) | 486.5 ( 209.3 ) | 645.8 ( 296.0 ) | <.0001 |

**Table 2.** Procedure Characteristics Stratified by Quartile of Annualized Hospital Volume in Atrial Fibrillation Ablation.

|  |  |  |  |  |  |  |
| --- | --- | --- | --- | --- | --- | --- |
| Characteristic | Overall | Quartile 1 | Quartile 2 | Quartile 3 | Quartile 4 | P value |
|  | (N = 70296) | (N = 5784) | (N = 11501) | (N =18597) | (N = 34414) |  |
| **Preprocedural imaging within prior 90 days** |  |  |  |  |  |  |
| Baseline transesophageal echocardiogram performed | 33533 (47.7%) | 3651 (63.1%) | 4475 (38.9%) | 9625 (51.8%) | 15782 (45.9%) | <.0001 |
| Baseline MRI performed | 2462 (3.5%) | 159 (2.7%) | 447 (3.9%) | 330 (1.8%) | 1526 (4.4%) | <.0001 |
| Baseline CT performed | 25580 (36.4%) | 2127 (36.8%) | 4192 (36.4%) | 7659 (41.2%) | 11602 (33.7%) | <.0001 |
| **Ablation strategy variables** |  |  |  |  |  |  |
| Phrenic nerve evaluation | 46252 (65.8%) | 3488 (60.3%) | 6253 (54.4%) | 12066 (64.9%) | 24445 (71.0%) | <.0001 |
| **Sedation** |  |  |  |  |  |  |
| Minimal sedation | 45 (0.1%) | 10 (0.2%) | 3 (0.0%) | 7 (0.0%) | 25 (0.1%) | 0.0013 |
| Moderate sedation | 528 (0.8%) | 82 (1.4%) | 122 (1.1%) | 137 (0.7%) | 187 (0.5%) | <.0001 |
| Deep sedation | 598 (0.9%) | 16 (0.3%) | 54 (0.5%) | 41 (0.2%) | 487 (1.4%) | <.0001 |
| General anesthesia | 69052 (98.2%) | 5663 (97.9%) | 11311 (98.3%) | 18386 (98.9%) | 33692 (97.9%) | <.0001 |
| **Isolation of all pulmonary veins (N=65,321)** | 64316 (98.5%) | 5141 (98.0%) | 10172 (98.0%) | 16830 (97.9%) | 32173 (98.9%) | <.0001 |
| Adjunctive ablation lesions | 37007 (52.6%) | 2836 (49.0%) | 5282 (45.9%) | 11124 (59.8%) | 17765 (51.6%) | <.0001 |
| Cavotricuspid isthmus ablation | 29073 (41.4%) | 2159 (37.3%) | 4118 (35.8%) | 8470 (45.5%) | 14326 (41.6%) | <.0001 |
| Cardioversion performed during procedure | 24712 (35.2%) | 1872 (32.4%) | 4007 (34.8%) | 7200 (38.7%) | 11633 (33.8%) | <.0001 |
| Atrial flutter/atrial tachycardia present during procedure | 22297 ( 31.7% ) | 1794 ( 31.0% ) | 3502 ( 30.4% ) | 5880 ( 31.6% ) | 11121 (32.3% ) | <.0001 |
| **Catheter manipulation** |  |  |  |  |  |  |
| Manual | 69935 ( 99.5% ) | 5745 ( 99.3% ) | 11454 (99.6% ) | 18425 (99.1% ) | 34311 (99.7% ) | <.0001 |
| Magnetic | 1823 ( 2.6% ) | 41 ( 0.7% ) | 403 ( 3.5% ) | 591 ( 3.2% ) | 788 ( 2.3% ) | <.0001 |
| Robotic | 153 ( 0.2% ) | 24 ( 0.4% ) | 23 ( 0.2% ) | 59 ( 0.3% ) | 47 ( 0.1% ) | <.0001 |
| **Procedural outcomes** |  |  |  |  |  |  |
| Radiation dose: Cumulative air kerma mGy, mean (std) | 278.6 ( 859.4 ) | 465.7(1100.7) | 250.6 ( 886.7 ) | 260.0 ( 949.8 ) | 269.8 ( 746.6 ) | <.0001 |
| (N=48,841) |  |  |  |  |  |  |
| **Discharge atrial rhythm (N=69,786)** |  |  |  |  |  |  |
| Sinus node rhythm | 66403 ( 94.5% ) | 5381 ( 93.0% ) | 10653 (92.6% ) | 17454 (93.9% ) | 32915 (95.6% ) | <.0001 |
| Atrial fibrillation | 1130 ( 1.6% ) | 126 ( 2.2% ) | 367 ( 3.2% ) | 294 ( 1.6% ) | 343 ( 1.0% ) | <.0001 |
| Atrial tachycardia | 136 ( 0.2% ) | 13 ( 0.2% ) | 34 ( 0.3% ) | 33 ( 0.2% ) | 56 ( 0.2% ) | 0.0377 |
| Atrial flutter | 211 ( 0.3% ) | 30 ( 0.5% ) | 52 ( 0.5% ) | 69 ( 0.4% ) | 60 ( 0.2% ) | <.0001 |
| Sinus arrest | 28 ( 0.0% ) | 5 ( 0.1% ) | 3 ( 0.0% ) | 15 ( 0.1% ) | 5 ( 0.0% ) | 0.0007 |
| Atrial paced | 2383 ( 3.4% ) | 199 ( 3.4% ) | 473 ( 4.1% ) | 674 ( 3.6% ) | 1037 ( 3.0% ) | <.0001 |
| **Discharge** |  |  |  |  |  |  |
| Home | 69898 (99.4%) | 5751 (99.4%) | 11433 (99.4%) | 18449 (99.2%) | 34265 (99.6%) | <.0001 |
| Other acute care hospital | 23 (0.0%) | 5 (0.1%) | 8 (0.1%) | 6 (0.0%) | 4 (0.0%) | 0.0022 |
| Left against medical advice | 33 ( 0.0% ) | 3 ( 0.1% ) | 6 ( 0.1% ) | 11 ( 0.1% ) | 13 ( 0.0% ) | 0.7282 |
| Discharged/transferred to an extended care/rehab | 110 ( 0.2% ) | 5 ( 0.1% ) | 13 ( 0.1% ) | 48 ( 0.3% ) | 44 ( 0.1% ) | 0.0006 |
| Skilled nursing facility | 138 ( 0.2% ) | 12 ( 0.2% ) | 23 ( 0.2% ) | 44 ( 0.2% ) | 59 ( 0.2% ) | 0.4451 |
| Other discharge location | 45 ( 0.1% ) | 2 ( 0.0% ) | 8 ( 0.1% ) | 27 ( 0.1% ) | 8 ( 0.0% ) | <.0001 |
| Discharge on any oral anticoagulant | 68695 (97.7%) | 5537 (95.7%) | 11190 (97.3%) | 18147 (97.6%) | 33821 (98.3%) | <.0001 |
| Discharge on antiarrhythmic drug | 36578 (52.0%) | 3422 (59.2%) | 6391 (55.6%) | 9522 (51.2%) | 17243 (50.1%) | <.0001 |

When quantified according to hospital procedural volume, patients in the lower quartiles were younger; had lower CHA_2_DS_2_VASc and HAS-BLED scores; and lower rates of comorbidities including heart failure, prior stroke, prior TIA, prior thromboembolic events, and vascular disease. Those in lower volume quartiles also had a higher mean BMI and were more likely to have persistent AF.

Similarly, for physician procedure volume quartiles **(Supplemental Table 1&2**), those in the lower quartiles were younger; were less commonly female; had lower CHA_2_DS_2_VASc score; and lower rate of comorbidities including heart failure, thromboembolic events, and vascular disease.

### Rates of Procedural Success and Adverse Events

The overall rate of acute procedural success was high at 98.5%. The rate of MAE was low (1%) and the rate of any AE was 2.2%. The most common AE were pericardial effusion requiring intervention (surgical or catheter, 0.4%), heart failure (0.4%), bradycardia (0.3%), respiratory failure (0.3%), hematoma at access site (0.2%), acute renal failure (0.1%), and TIA or stroke (0.1%).

### Annual Hospital volume and outcomes

The median hospital procedure volume was 230 (IQR: 144.7-307.2). The median number of procedures across the quartiles were Q1: 66 (IQR: 53.5-76.7), Q2: 115.2 (IQR: 102.2-128), Q3: 197 (IQR: 167.2-208.3) and Q4: 307.2 (IQR: 280-433). The procedural success rates were high overall across hospital volume quartiles, but improved with increasing hospital volume at 98.0% (Q1), 98.0% (Q2), 97.9% (Q3) and 99.0% (Q4) (**Table 3**). When compared with Q4 (highest quartile) as the reference, the adjusted odds of procedural success were lower in Q1 (OR: 0.44, CI: 0.29-0.68, p=0.0002), Q2 (OR: 0.50, CI: 0.33-0.75, p=0.001), and Q3 (OR: 0.60, CI: 0.40-0.89, p=0.011) as shown in **Figure 2A**. **Figure 3A** depicts the unadjusted and adjusted relationship between hospital volume and procedural success. An adjusted procedural success of 98% was predicted with an annual hospital volume of approximately 154 or more procedures as depicted in **Figure 3A (dotted orange line**).

**Figure 2:**
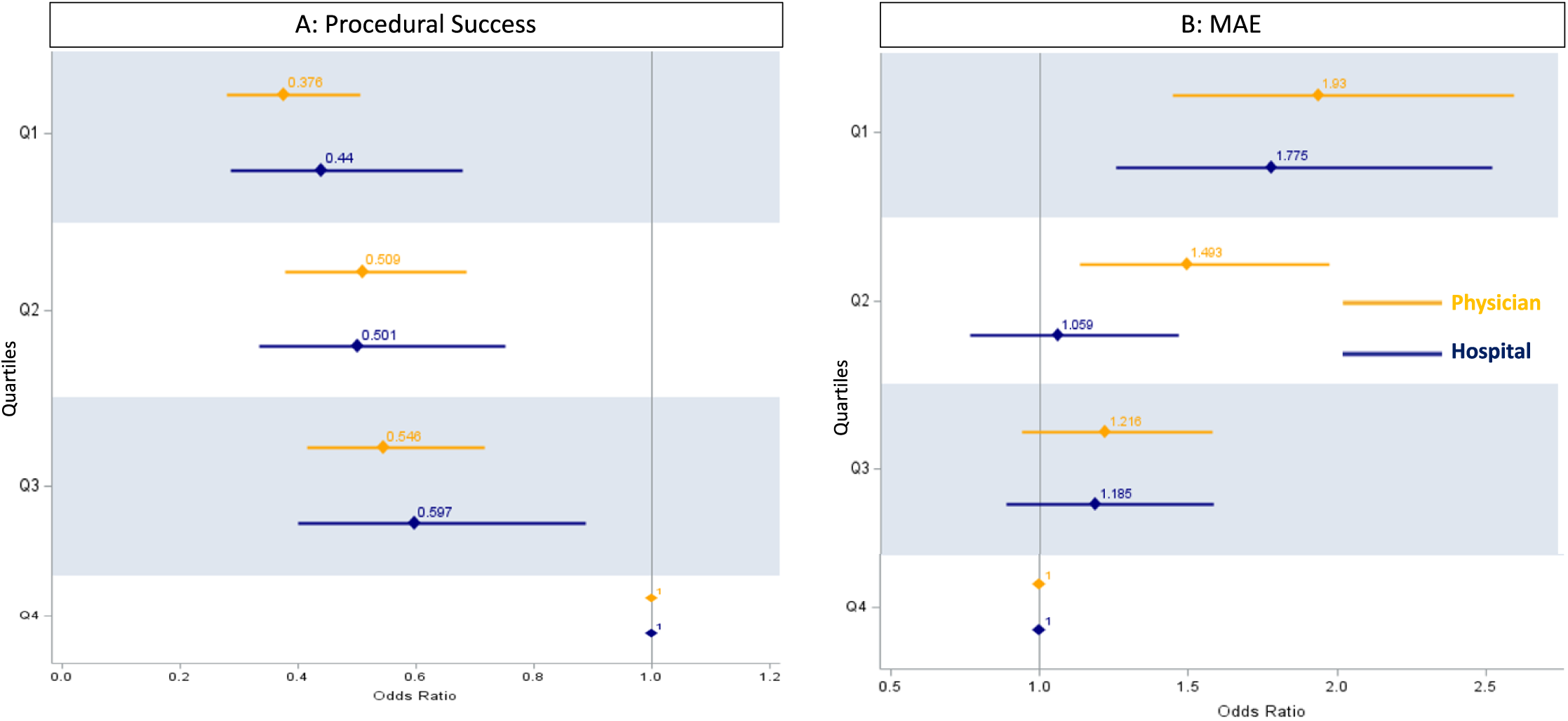
Annual Procedure Volume, Procedural Success, and MAE. Adjusted relationship between annual hospital/physician volume quartile (Q) and procedural success [panel A] and MAE [panel B] respectively when compared to Q4 (highest volume quartile). Panel A shows lower odds of procedural success in Q1, Q2, and Q3 for both hospital and physician compared to Q4. Panel B shows higher odds of MAE in Q1 for both hospital and physician when compared to Q4. Procedural success, defined by isolation of all pulmonary veins; MAE, Major Adverse Events; Q, Procedure volume quartiles

**Figure 3:**
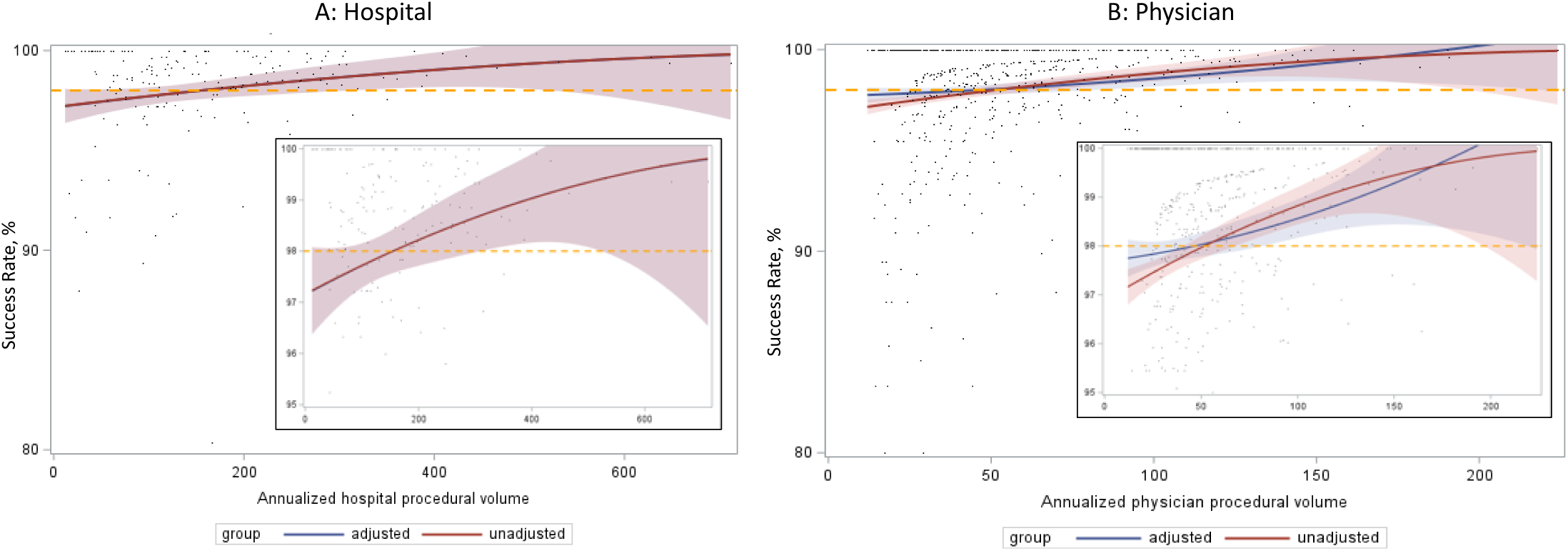
Hospital and Physician Procedural Volume and Procedural Success. Relationship between procedural success and annualized hospital (Panel A) and physician (Panel B) procedural volume. There is a positive correlation between procedural volume and procedural success for both hospitals and physicians. Dotted orange line depicts an adjusted procedural success of ≥98%, that can be achieved with an annual hospital and physician volume of approximately ≥150 (Panel A) and ≥50 (Panel B) cases, respectively. Procedural success, defined by isolation of all pulmonary veins

**Table 3.** Incidence of Adverse Events Following AF Ablation Stratified by Quartile of Annualized Hospital Volume.

| Outcomes | Overall | Quartile 1 | Quartile 2 | Quartile 3 | Quartile 4 | P value |
| --- | --- | --- | --- | --- | --- | --- |
|  | (N =70296) | (N =5784) | (N =11501) | (N =18597) | (N =34414) |  |
| Major adverse event* | 702 ( 1.0% ) | 82 ( 1.4% ) | 116 ( 1.0% ) | 191 (1.0% ) | 313 ( 0.9% ) | 0.0042 |
| In-hospital death | 2 ( 0.0% ) | 0 ( 0.0% ) | 1 ( 0.0% ) | 0 ( 0.0% ) | 1 ( 0.0% ) | 0.7385 |
| Hospitalization >1 day | 6572 ( 9.3% ) | 626 ( 10.8% ) | 1010 ( 8.8% ) | 1853 ( 10.0% ) | 3083 ( 9.0% ) | <.0001 |
| Same day Discharge | 28277 ( 40.2% ) | 1715 ( 29.7% ) | 4396 ( 38.2% ) | 7412 ( 39.9% ) | 14754 ( 42.9% ) | <.0001 |
| Any adverse event | 1539 ( 2.2% ) | 156 ( 2.7% ) | 272 ( 2.4% ) | 441 ( 2.4% ) | 670 ( 1.9% ) | 0.0001 |
| **Specific adverse events** |  |  |  |  |  |  |
| Bradycardia adverse events | 235 ( 0.3% ) | 23 ( 0.4% ) | 44 ( 0.4% ) | 77 ( 0.4% ) | 91 ( 0.3% ) | <.0001 |
| Cardiac arrest | 67 ( 0.1% ) | 6 ( 0.1% ) | 14 ( 0.1% ) | 23 ( 0.1% ) | 24 ( 0.1% ) | 0.001 |
| Myocardial infarction | 20 ( 0.0% ) | 3 ( 0.1% ) | 6 ( 0.1% ) | 4 ( 0.0% ) | 7 ( 0.0% ) | 0.0007 |
| Air embolism | 16 ( 0.0% ) | 3 ( 0.1% ) | 3 ( 0.0% ) | 4 ( 0.0% ) | 6 ( 0.0% ) | 0.0036 |
| LA thrombus | 3 ( 0.0% ) | 1 ( 0.0% ) | 0 ( 0.0% ) | 1 ( 0.0% ) | 1 ( 0.0% ) | 0.0032 |
| Cardiac thromboembolic event | 5 ( 0.0% ) | 0 ( 0.0% ) | 0 ( 0.0% ) | 3 ( 0.0% ) | 2 ( 0.0% ) | 0.0026 |
| Stroke | 77 ( 0.1% ) | 7 ( 0.1% ) | 16 ( 0.1% ) | 22 ( 0.1% ) | 32 ( 0.1% ) | 0.0031 |
| TIA | 16 ( 0.0% ) | 1 ( 0.0% ) | 3 ( 0.0% ) | 3 ( 0.0% ) | 9 ( 0.0% ) | 0.0053 |
| TIA or stroke | 93 ( 0.1% ) | 8 ( 0.1% ) | 19 ( 0.2% ) | 25 ( 0.1% ) | 41 ( 0.1% ) | 0.6914 |
| Arterial thrombosis | 7 ( 0.0% ) | 1 ( 0.0% ) | 0 ( 0.0% ) | 5 ( 0.0% ) | 1 ( 0.0% ) | 0.0003 |
| Deep vein thrombosis | 11 ( 0.0% ) | 2 ( 0.0% ) | 3 ( 0.0% ) | 2 ( 0.0% ) | 4 ( 0.0% ) | 0.0035 |
| Respiratory failure | 222 ( 0.3% ) | 16 ( 0.3% ) | 44 ( 0.4% ) | 50 ( 0.3% ) | 112 ( 0.3% ) | 0.0018 |
| Phrenic nerve damage | 86 ( 0.1% ) | 16 ( 0.3% ) | 7 ( 0.1% ) | 22 ( 0.1% ) | 41 ( 0.1% ) | <.0001 |
| Pulmonary embolism | 8 ( 0.0% ) | 2 ( 0.0% ) | 1 ( 0.0% ) | 1 ( 0.0% ) | 4 ( 0.0% ) | 0.0027 |
| Pulmonary vein damage/dissection | 3 ( 0.0% ) | 0 ( 0.0% ) | 0 ( 0.0% ) | 0 ( 0.0% ) | 3 ( 0.0% ) | 0.003 |
| Pneumonia | 64 ( 0.1% ) | 4 ( 0.1% ) | 14 ( 0.1% ) | 21 ( 0.1% ) | 25 ( 0.1% ) | 0.0023 |
| Sepsis | 9 ( 0.0% ) | 2 ( 0.0% ) | 2 ( 0.0% ) | 2 ( 0.0% ) | 3 ( 0.0% ) | 0.0034 |
| Acute renal failure | 102 ( 0.1% ) | 5 ( 0.1% ) | 23 ( 0.2% ) | 24 ( 0.1% ) | 50 ( 0.1% ) | 0.002 |
| GU bleeding | 27 ( 0.0% ) | 3 ( 0.1% ) | 5 ( 0.0% ) | 8 ( 0.0% ) | 11 ( 0.0% ) | 0.005 |
| Heart failure | 268 ( 0.4% ) | 11 ( 0.2% ) | 54 ( 0.5% ) | 77 ( 0.4% ) | 126 ( 0.4% ) | 0.0003 |
| Pericardial effusion resulting in cardiac tamponade | 168 ( 0.2% ) | 21 ( 0.4% ) | 25 ( 0.2% ) | 51 ( 0.3% ) | 71 ( 0.2% ) | 0.0007 |
| Pericardial effusion requiring intervention (pericardiocentesis or surgical intervention) | 297 ( 0.4% ) | 37 ( 0.6% ) | 48 ( 0.4% ) | 76 ( 0.4% ) | 136 ( 0.4% ) | 0.0006 |
| Cardiac surgery | 48 ( 0.1% ) | 7 ( 0.1% ) | 9 ( 0.1% ) | 9 ( 0.0% ) | 23 ( 0.1% ) | 0.0016 |
| Hemorrhage (non-access site) | 99 ( 0.1% ) | 6 ( 0.1% ) | 20 ( 0.2% ) | 23 ( 0.1% ) | 50 ( 0.1% ) | 0.0049 |
| Hematoma at access site | 154 ( 0.2% ) | 19 ( 0.3% ) | 27 ( 0.2% ) | 42 ( 0.2% ) | 66 ( 0.2% ) | 0.0017 |
| Bleeding requiring transfusion (access site) | 61 ( 0.1% ) | 7 ( 0.1% ) | 13 ( 0.1% ) | 17 ( 0.1% ) | 24 ( 0.1% ) | 0.0033 |
| AV fistula requiring intervention | 4 ( 0.0% ) | 3 ( 0.1% ) | 0 ( 0.0% ) | 1 ( 0.0% ) | 0 ( 0.0% ) | <.0001 |
| Pseudoaneurysm requiring intervention | 71 ( 0.1% ) | 4 ( 0.1% ) | 16 ( 0.1% ) | 19 ( 0.1% ) | 32 ( 0.1% ) | 0.0039 |
| *Major adverse event was defined as a composite of death, all-cause stroke, transient ischemic attack, cardiac arrest, acute myocardial infraction, pericardial effusion requiring intervention, bleeding requiring transfusion, and vascular injury requiring intervention. |  |  |  |  |  |  |

The MAE rate was low across all volume quartiles, but decreased with increasing hospital volume at 1.4% (Q1), 1.0% (Q2), 1.0% (Q3), and 0.9% (Q4). When compared with Q4, the adjusted odds of a MAE were significantly higher for Q1 (OR: 1.78, CI: 1.26-2.52, p=0.001) but not for Q2 (OR: 1.06, CI: 0.77-1.46, p=0.728) or Q3 (OR: 1.19, CI: 0.89-1.58, p=0.249) as depicted in **Figure 2B**. **Figure 4A** depicts the unadjusted and adjusted relationship between hospital volume and procedural safety. An adjusted procedural MAE of less than 1% was predicted by an annual hospital volume of approximately 190 or more procedures as depicted in **Figure 4A (dotted orange line).** There appears to be a ceiling effect with a slight increase in AE rates for the highest volume centers, although the number of centers with very high volumes was low and the confidence intervals were markedly wider at the upper extreme of volume.

**Figure 4:**
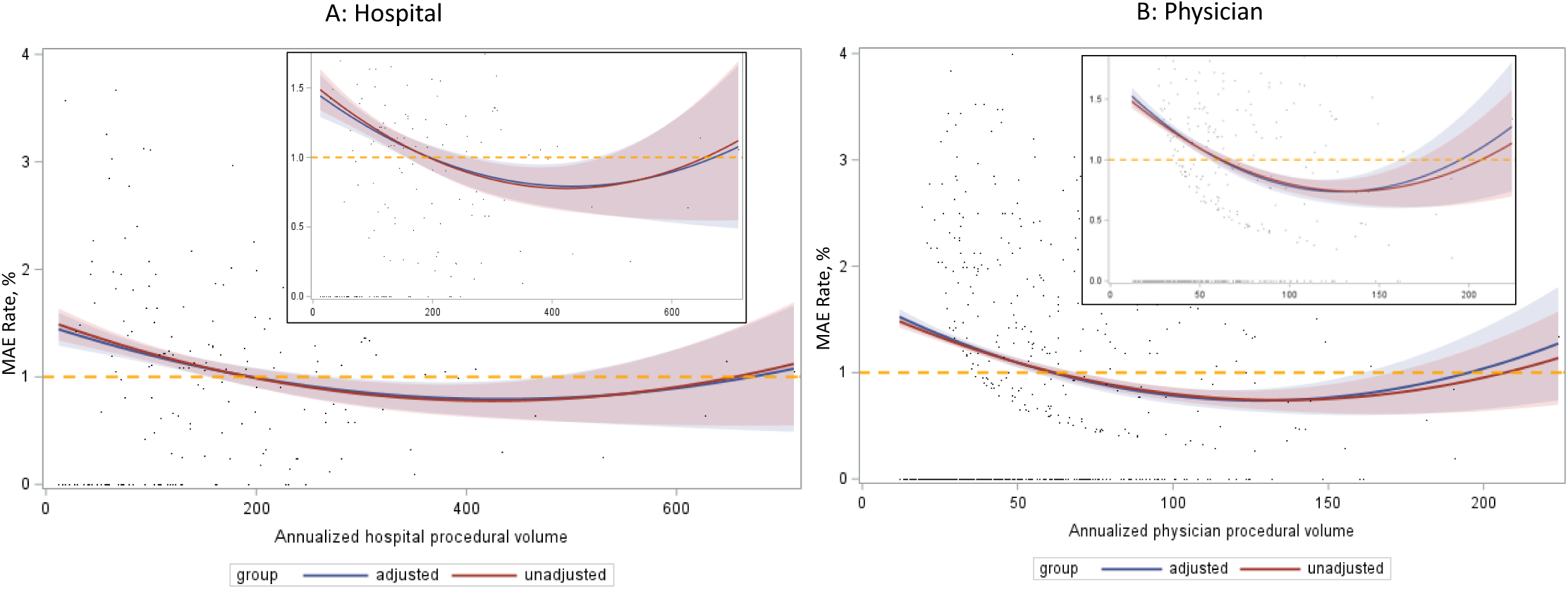
Hospital and Physician Procedural Volume and MAE. Relationship between MAE and annualized hospital (Panel A) and physician (Panel B) procedural volume. There is an inverse relationship between MAE and procedural volume for both hospitals and physicians. Dotted orange line depicts an adjusted procedural MAE ≤1% that can be achieved with an annual hospital and physician volume of approximately ≥190 and ≥60 cases, respectively. MAE, Major Adverse Events;

**Figure 5:**
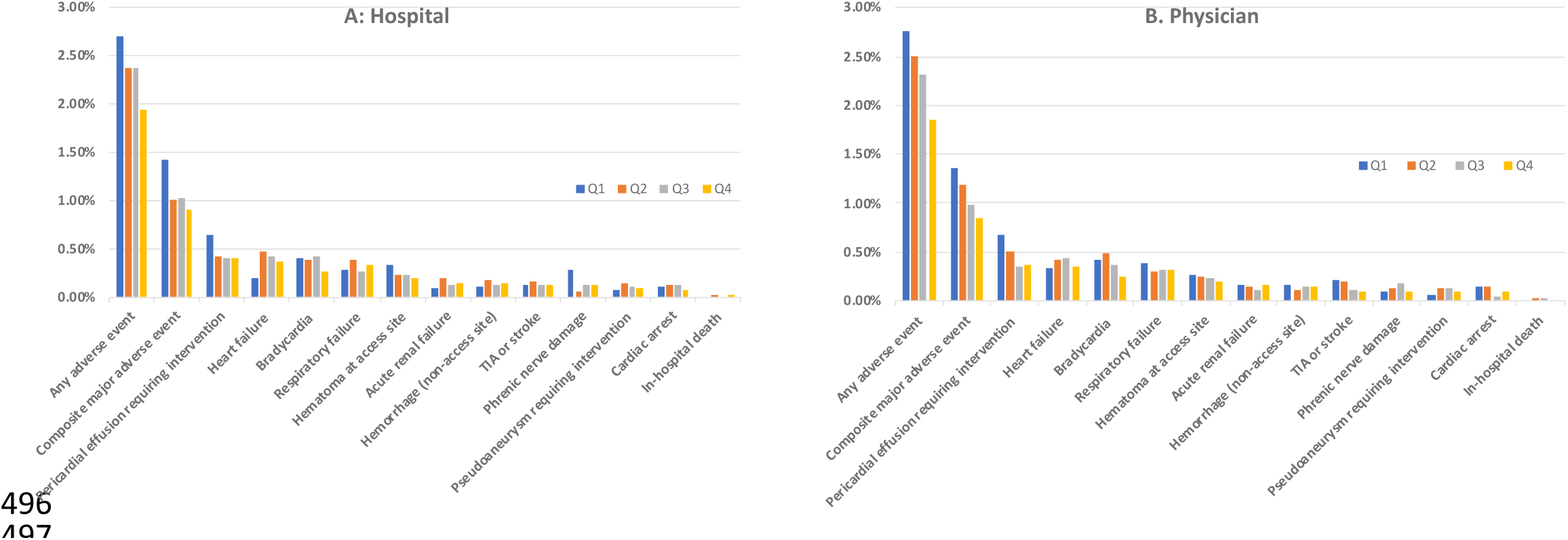
Adverse Events Stratified by Annualized Hospital and Physician Volume. Incidence of adverse events stratified by annualized hospital (Panel A) and physician (Panel B) procedure volume quartiles Q, Procedure Volume Quartiles

The overall rate of any AE was 2.2% and decreased with increasing hospital procedural volume at 2.7%(Q1), 2.4% (Q2), 2.4% (Q3) and 1.9%(Q4), (P<0.0001) respectively. When compared with Q4, the adjusted odds of any AE were higher for Q1 (OR: 1.51, CI: 1.12-2.02, p=0.007) but not for Q2 (OR: 1.15, CI: 0.88-1.51, p=0.299) and Q3 (OR: 1.24, CI: 0.96-1.63, p=0.093) **(Supplemental Figure 1)**.

Patients who had their procedure performed in higher volume hospitals had a higher rate of same day discharge, Q1(29.7%), Q2 (38.2%), Q3(39.9%), and Q4 (42.9%), respectively.

### Annual physician volume and outcomes

The median annual physician procedure volume was 66 (IQR: 44.7-97.5). The median number of procedures across the quartiles were Q1: 26.9 (IQR: 23.5-30), Q2: 40.2 (IQR: 36.9-44), Q3: 60 (IQR: 54-64.3), and Q4: 103 (IQR: 85.9-126). The procedural success rate across the quartiles was high, but increased across volume quartiles at 97.6% (Q1), 98.1% (Q2), 97.9% (Q3), and Q4 99.1% (Q4) (**Table 4**). When compared with Q4 as the reference, the adjusted odds of procedural success were lower in Q1 (OR: 0.38, CI: 0.28-0.51, p<0.001), Q2 (OR: 0.51, CI: 0.38-0.69, p<0.001), and Q3 (OR: 0.55, CI: 0.42-0.72, p<0.001) as shown in **Figure 2A**. **Figure 3B** depicts the unadjusted and adjusted relationship between annual physician volume and procedural success. An adjusted procedural success of 98% was predicted by an annual physician volume of approximately 50 or more procedures as depicted in **Figure 3B (dotted orange line)**

**Table 4.**
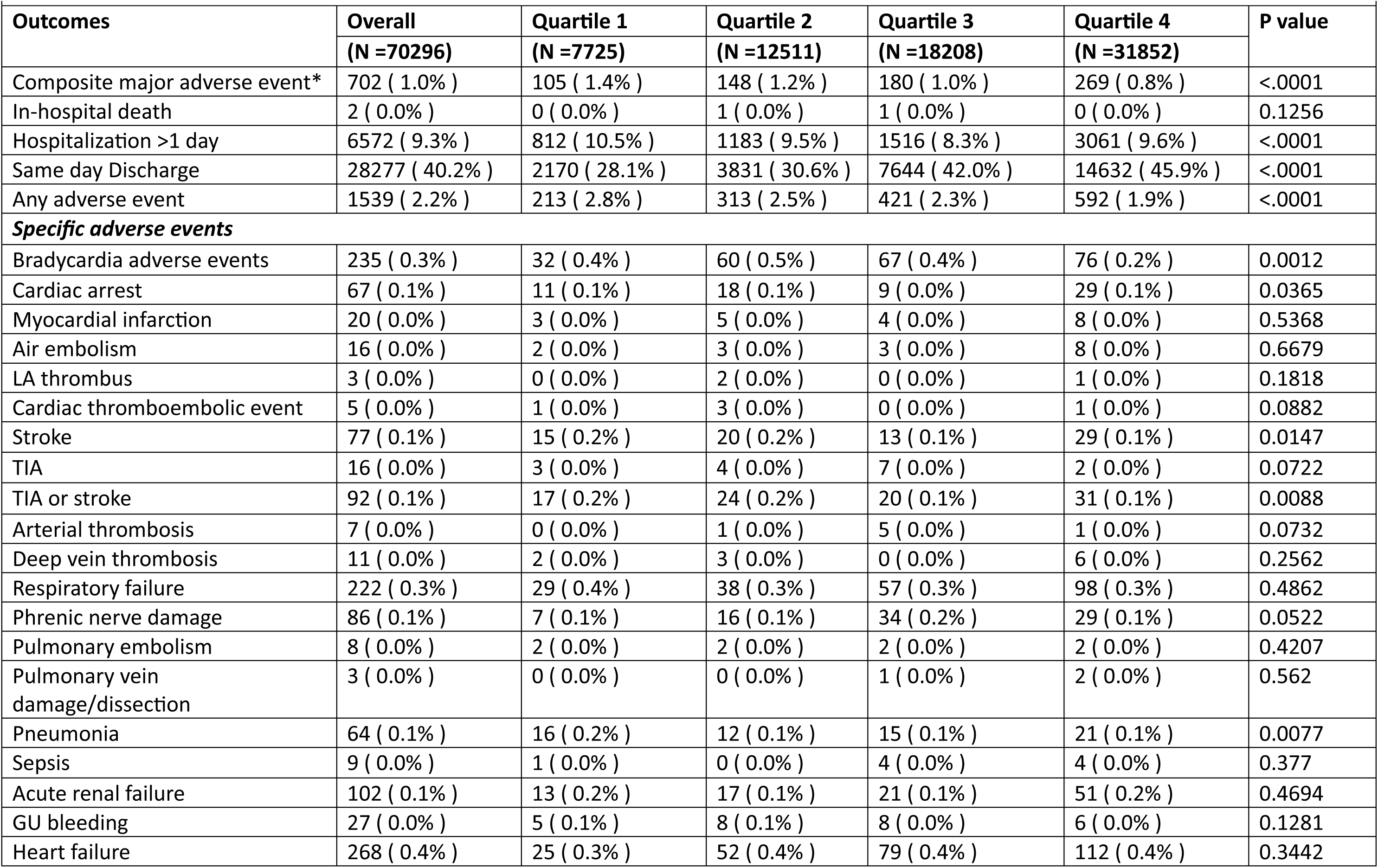
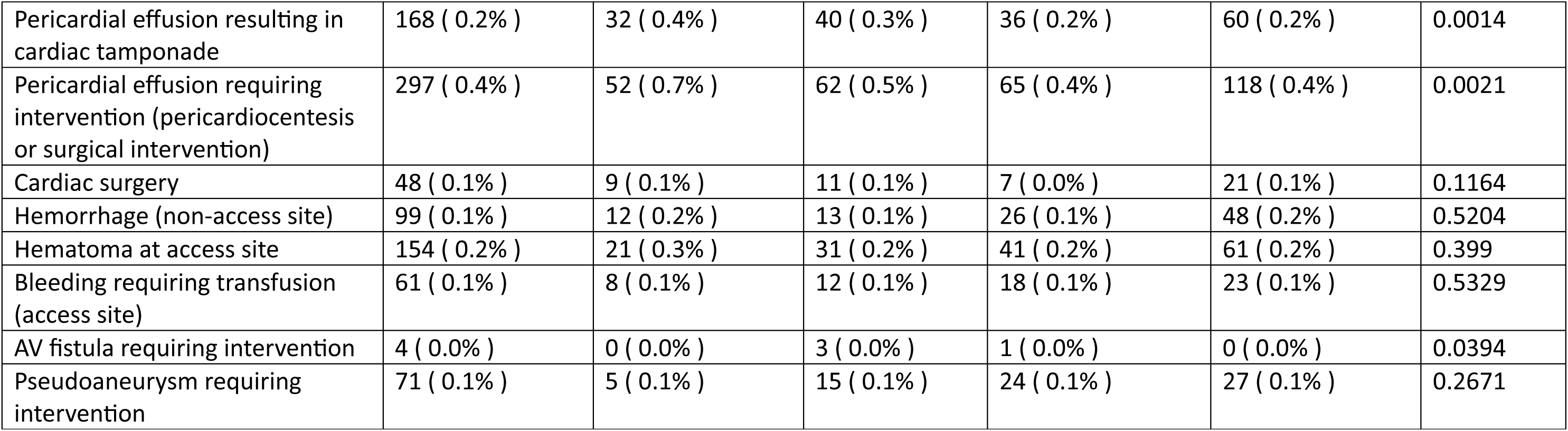
Incidence of Adverse Events Following AF Ablation Stratified by Quartile of Annualized Physician Volume.

The procedural MAE rate across the physician volume quartiles were 1.4% (Q1), 1.2% (Q2), 1.0% (Q3), and 0.8% (Q4). When compared with Q4, the odds of an MAE were higher in Q1 (OR: 1.93, CI: 1.44-2.58, p<0.0001) and Q2 (OR: 1.49, CI: 1.13-1.97, p=0.004), but not in Q3 (OR: 1.22, CI: 0.94-1.58, p=0.140) as depicted in **Figure 2B**. **Figure 4B** depicts the unadjusted and adjusted relationship between physician volume and safety. An adjusted procedural MAE of less than 1% was predicted by an annual physician volume of approximately 62 or more procedures as depicted in **Figure 4B (dotted orange line).** There appears to be a ceiling effect with a slight increase in AE rate for the highest volume physicians, although the number of physicians with very high volumes was low and the confidence intervals were substantially wider at the upper extreme of volume.

Patients who had their procedure performed by physicians in a higher volume quartile had higher likelihood of same day discharge, Q1 (28.1%), Q2 (30.6%), Q3 (42.0%), and Q4 (45.9%), respectively.

### Sensitivity analysis not restricting to first time AF ablation procedures

In a secondary analysis including all patients who had undergone AF ablation, not restricting to first time AF ablations only, there were a total of 85,585 patients from 187 hospitals across US. In this cohort, the findings were consistent with the results of the primary analysis. There was a significant inverse relationship between both physician and hospital procedural volume and MAE rates **(Supplemental Tables 3 & 4 and Supplemental Figures 2A & 2B)**.

### Secondary adjusted analysis adjusting for total procedural volume

In a secondary analysis adjusting for the total procedural volume performed by a hospital during the study period, the odds of procedural success were lower in lower quartiles when compared with Q4, consistent with our primary analysis. However, for safety endpoint (MAE & any AE), after adjustment for total hospital procedure volume, it lost its statistical significance **(Supplemental Figures 3A & 3B and 4).**

Similarly, for physicians, with adjustment for total procedures performed during the study period, the odds of procedural success were lower and rate of MAE or any AE were higher in lower quartiles when compared to Q4, consistent with our primary analysis **(Supplemental Figures 3A & 3B and 4).**

## Discussion

In this large cohort of over 70,000 AF ablations from 186 US hospitals treated by 753 physicians, we uncovered several important findings. First, the overall rate of acute procedural success was high at 98.5% and the rates of major and any AE were relatively low at 1% and 2.2%, respectively. Second, increasing hospital and physician volumes were associated with significantly higher rates of acute procedural success and lower rates of MAEs. This relationship was particularly strong for physicians. We found that an adjusted procedural MAE rate of less than 1% is predicted by an approximate annual volume of 60 for physicians and 190 for hospitals.

This study highlights the overall improved acute procedural success and safety of AF ablation in contemporary US practice. The rates of procedural complications in this study were low compared with prior published data and in-hospital death was very rare, with only 2 patients in over 70,000 procedures. A recent study looking for 30-day mortality in over 122,000 Medicare patients between 2016-2019, showed overall mortality of 0.6 % and post-procedure complication rate of 6.5%, higher than in our study but the participants were limited to Medicare beneficiaries who are much older and have higher comorbidities than average participants undergoing catheter ablation ^17^. Earlier data from the NIS from 2000-2017 also showed higher rates of AE than we found in our study, with AE in 5.4-7.2% and in-hospital mortality of 0.15%-0.46% ^11,18,19^. However, it is vital to note that NIS data only capture inpatient hospitalization status procedures, many of which resulted in post-procedure admissions because the patient experienced a complication or underwent ablation during an acute hospitalization. As such, these results are not generalizable to the majority of contemporary AF ablations which are conducted with same day discharge or an outpatient observation hospitalization status. Similarly, nationwide administrative data for AF ablation in 2014 from Germany showed higher complication rates up to 13.8% and major complication rates up to 7.2% ^20^. More recently, the prospective CABANA trial also reported the overall complication rate of 5.9% in patients who underwent AF ablation ^21^. Besides taking into account ambulatory procedures, the lower rate of complications noted in our study from the NCDR AFib Ablation Registry compared with earlier studies may be partially explained by variation in the definitions of complications, continued improvements in catheter design including contact force sensing, improved peri-procedural management including common use of uninterrupted anticoagulation, more widespread use of electroanatomic mapping systems, and increased use of intracardiac echocardiography. In addition, the AFib Ablation Registry does not capture the AE that occur after the index procedure hospitalization so procedure related AE occurring after discharge are not captured. In addition, registry participation is voluntary, with a focus on quality improvement initiatives, so findings may not be fully generalizable to non-participating centers. Finally, underreporting is possible in this voluntary registry, although the NCDR used a rigorous data quality reporting process that includes audits of adverse events at 5% of sites annually using manual review of medical records and billing codes.

The association between procedure volume and outcomes has been established for other cardiovascular procedures including transcatheter aortic valve replacement ^10^, implantable cardioverter defibrillator implantation ^9^, percutaneous coronary intervention ^8^, coronary artery bypass surgery^5^, and mitral valve surgery ^6^. In this study, we observed a significant relationship between hospital and physician procedure volume and acute procedural success and safety. A secondary analysis that included both first-time and re-do AF ablation showed consistent results. These findings corroborate and expand upon prior studies that were older and limited by the study populations and methods used. An NIS study including 14,875 inpatient AF ablation from 2015-2017 showed significantly higher overall complication rates in low-volume centers compared with high centers ((12.15 vs 4.75%, p<.001; high volume defined as >115 procedures/year) and a non-significant difference in mortality (0.63% vs. 0.22%, p=0.244)^19^. Similarly, a 2014 using administrative data of AF ablations from Germany showed a significantly higher rate of overall complications in low-volume centers compared with high volume centers (16.4% vs 12.7%, p=0.0002; high volume defined as >100/year), but no difference in major complications ^20^. A recent meta-analysis on AF ablation procedure volume and outcomes showed high volume hospitals and physicians (≥100 procedures/ year) had lower complication rates [5.5% vs. 6.2%, p<0.001; OR 0.62 (0.53-0.73)] and [3.69% vs. 7.25%, p<0.01; OR=0.49 (0.30-0.80)], respectively ^22^. These studies were limited by the use of relatively arbitrary cut-offs to define low vs high volume centers, and the use of inpatient administrative claims data for several studies that are likely to overestimate complication rates. Recently published data from Japan’s national database of healthcare insurance claims from 2014 to 2020 involving more than 270,000 de-novo AF ablations, showed an acute complication rate of 2%, similar to our study and acute procedural complications decreased significantly when hospital procedural volume approached 150-200/year. Interestingly, this volume-outcome effect was seen only for radiofrequency ablation, but not for cryoablation ^23^. In our study, we did not stratify by ablation energy source as this was not an available variable in the registry, but this attenuated volume-outcome relationship with cryoablation may be due to less procedural and patient complexity and a potentially faster learning curve for hospital staff and physicians. As more ablation technologies for PVI such as pulsed field ablation are commercially released ^24^, the volume-outcome relationship may change.

Further, in our cohort, the procedural success above 98% was predicted by annual procedure volumes at the hospital and physician level of 154 and 50 cases, respectively. Similarly, we found that a MAE rate below 1% was predicted by an annual case volume at the hospital and physician level of 194 and 62, respectively. There appears to be a ceiling effect and a slight increase in adverse event rates after achieving certain procedure volume for both hospitals and physicians, although the number of centers with very high volumes was low and the confidence intervals for MAEs were very wide at the upper extreme of volume. It is possible that high-volume centers and physicians are willing to perform procedures on higher risk patients and there is some residual unmeasured confounding that our adjustment cannot account for with these higher risk patients. Adjusting for cumulative procedure volume in general did not change the association between annualized procedure volume and outcomes with the exception that at high cumulative hospital volume did seem to attenuate the effect somewhat.

The optimal procedural volumes for hospitals and physicians are not clear, and various studies have suggested volume thresholds for hospitals and physicians of 100-200 and 25-100 per year, respectively^11,12,17,19,20,22,23^. Based on our current study, hospitals and physicians performing approximately 190 and 60 annual cases, respectively, are likely to contribute to consistently high procedural success (>98%) and safety (MAE <1%). In our cohort, the nadir of MAE rates occurred for hospitals at a volume of approximately 400 procedures annually and for physicians with an annual volume of approximately 120 procedures, but the improvements in risk above the previously defined thresholds were modest. Taken together, these findings suggest that moderate procedural volumes that are likely achievable across a variety of healthcare systems may be sufficient to maintain the necessary skills of hospital staff and physicians to achieve favorable procedural success and safety. These findings may inform future AF ablation guideline recommendations ^1^, hospital and physician practice quality improvement initiatives and establish criteria for AF centers of excellence as was recently proposed in a 2020 white paper from the Heart Rhythm Society ^25^.

## Limitations

This study has several limitations. With any registry-based study, where patient and procedural characteristics are site-reported, underreporting of adverse events or overstating procedural success may be possible. However, the NCDR registries uses a data quality control program to ensure that submissions are valid, complete and accurate with annual audits of 5% of the participating sites ^15^. The data for this study reflect the characteristics of participating hospitals in the NCDR AFib Ablation Registry that may not be completely generalizable. The period of study included the major worldwide pandemic from COVID-19 which may have affected the procedures performed by the individual physicians or hospital systems. However, the study includes data from over 70,000 patients at 186 sites, including during periods before and after the peak of the pandemic, and the AFib Ablation Registry is the largest prospective registry of AF ablation procedures worldwide at present. Finally, individual physician training and duration in practice performing procedures may also affect procedural outcomes, but these data are not currently available in the registry. However, we did a secondary analysis adjusting for the total procedure volume performed by a hospital or physician during the study period and these results were overall very consistent with the results of our primary analyses.

## Conclusion

In this national cohort of first time AF ablations, acute procedural success was relatively high and the rate of AE was low. Higher hospital and physician procedure volume was significantly associated with increased acute procedural success and lower risk of AE. An adjusted procedural MAE rate of less than 1% is predicted by an annual volume of approximately 190 for hospital and 60 for physicians. These findings may inform guideline recommendations regarding minimum procedural volumes for AF ablation.

## Perspectives

### What is new?

- Our study shows, in contemporary practice, the acute procedural success in atrial fibrillation ablation is high and rate of major adverse events are low.
- Higher hospital and physician procedure volume is associated with greater acute procedural success and lower risk of adverse events

### What are the clinical implications?

- These findings may help guideline recommendation on procedural volume, for both hospital and physician, to maintain procedural competency to achieve high procedural success and low adverse event in atrial fibrillation ablation.

## Data Availability

The data for the study were obtained from the NCDR AF registry. Data may be made available from NCDR AF registry upon request.

## Abbreviation lists

AF: Atrial fibrillation
MAE: Major adverse events
AE: Adverse events
NCDR: National Cardiovascular Data Registry
Q: Quartiles
CORE: Center for Outcomes Research and Evaluation
NIS: National Inpatient Sample
GFR: Glomerular filtration rate
CHF: Congestive heart failure
TIA: Transient ischemic attack
BMI: Body mass index
IQR: Interquartile range

---

**Figure:**
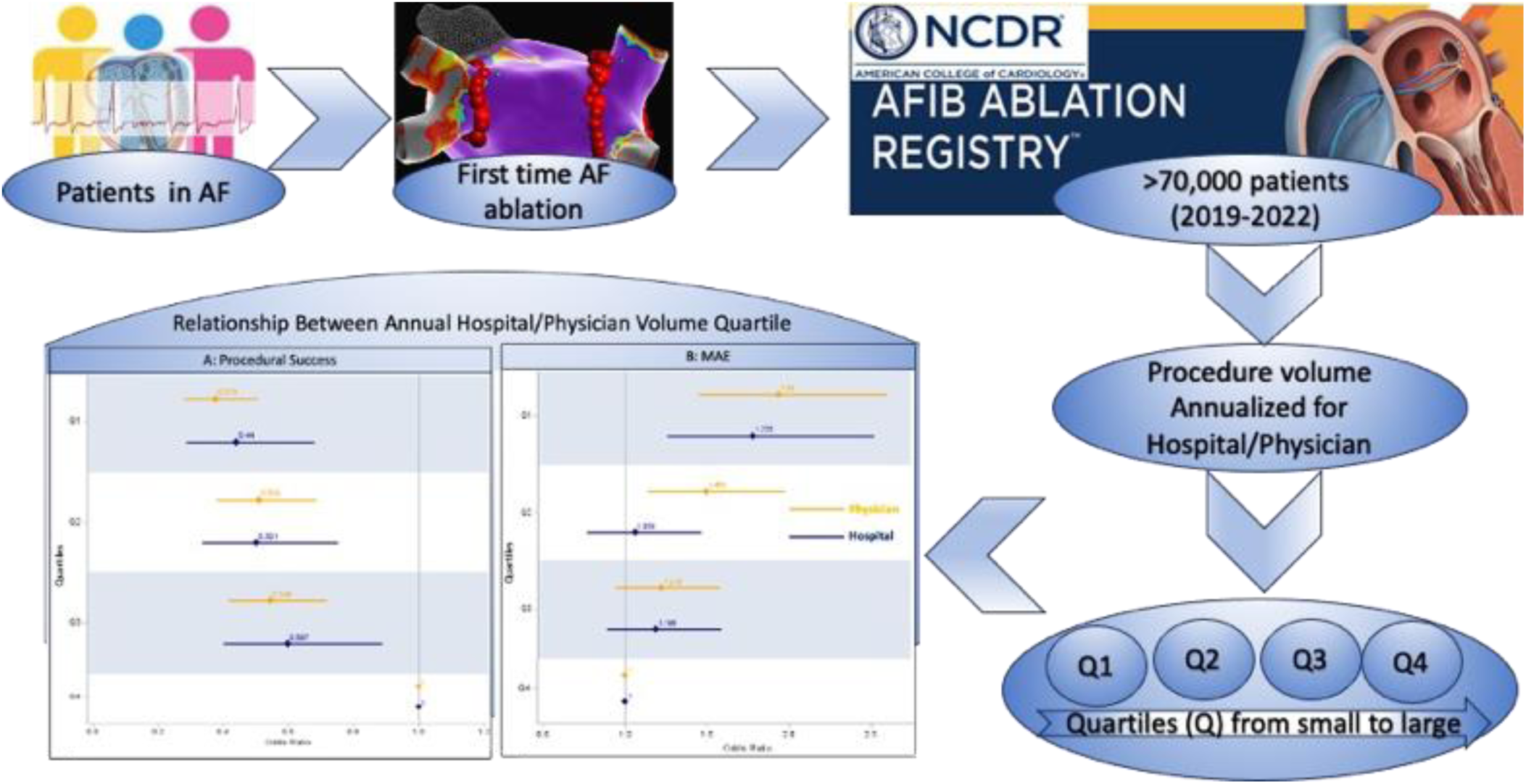
Central Illustration. Patients enrolled in NCDR AF ablation registry between July 2019-June 2022 were categorized into procedure volume quartiles and compared for procedural success (defined by isolation of all pulmonary veins) and adverse events AF, Atrial fibrillation; NCDR, National Cardiovascular Data Registry; Q, Quartiles; MAE, Major adverse events

